# Maternal health around pregnancy and autism risk: a population-based study

**DOI:** 10.1101/2020.05.19.20089581

**Authors:** Arad Kodesh, Stephen Z. Levine, Vahe Khachadourian, Rayees Rahman, Avner Schlessinger, Paul F. O’Reilly, Jakob Grove, Diana Schendel, Joseph D. Buxbaum, Lisa Croen, Abraham Reichenberg, Sven Sandin, Magdalena Janecka

## Abstract

**IMPORTANCE:** Many maternal diagnoses in pregnancy have been linked with offspring ASD risk. However, pregnant women receive many other diagnoses, most of which have not been evaluated for an association with offspring ASD.

**OBJECTIVE:** Systematically test the associations between maternal diagnoses around pregnancy, and ASD risk in offspring, filtering out potential false positives and accounting for correlation between maternal diagnoses.

**DESIGN, SETTING AND PARTICIPANTS:** This case-cohort study included children born in Israel from January 1, 1999, through December 31, 2008, and followed up until January 26, 2015. We used information on all ICD-9 codes received by their mothers during pregnancy and the preceding year. ASD risk in children whose mothers experienced each of those conditions during the exposure period was compared with ASD risk in unexposed children. Data were analyzed from December 1, 2019, through April 30, 2020.

**MAIN OUTCOME AND MEASURES:** Hazard ratios (HRs) and 95% confidence intervals (CIs) of ASD risk associated with each maternal ICD-9 code, calculated using Cox proportional hazards regression, adjusted for the confounders (birth year, maternal age, socioeconomic status and number of ICD-9 diagnoses during the exposure period).

**RESULTS:** The analytic sample consisted of 80,187 individuals (1,132 cases, 79,055 controls; median [interquartile range] age at the end of follow up was 11.2 [8.8-13.6]; 48.8% female). Of 822 ICD-9 codes tested, 148 were recorded in at least 10 of each case and control mothers. Of those, 22 maternal diagnoses were nominally significantly associated with offspring ASD, and 16 of those survived subsequent filtering steps (permutation testing, multiple testing correction, multiple regression). Among those 16 diagnoses, we recorded increased risk of ASD associated with certain metabolic (e.g. hypertension (ICD-9 401); HR=2.74(1.92-3.90), P=2.43E^-8^), genitourinary (e.g. noninflammatory disorders of cervix (ICD-9 622); HR=1.88(1.38-2.57), P=7.06E^-5^) and psychiatric (depressive disorder (ICD-9 311); HR=2.11(1.32-3.35), P=1.70E^-3^) diagnoses. Meanwhile, mothers of children with ASD were less likely to attend prenatal care appointment (ICD-9 V22; HR=0.62 (0.54-0.71), P=1.80E^-11^).

**CONCLUSIONS AND RELEVANCE:** We observed an association between ASD and 16 maternal ICD-9 diagnoses, after rigorous filtering out potential false positive associations. Replication in other cohorts and further research to understand the mechanisms underlying the observed associations with ASD are warranted.

## INTRODUCTION

ASD is a neurodevelopmental disorder affecting 1.7% of children in the US^1^, and characterized by difficulties in social communication and rigid, repetitive behaviors. While the disorder likely arises due to a combination of genetic and environmental factors^2^, potential modifiable risk factors for this disorder remain unknown.

As the pathology underlying ASD likely originates *in utero*, search of the actionable, etiological risk factors for the disorder has focused on maternal exposures in pregnancy. Although the fetus is protected from many such exposures due to the placental barrier - which ensures lack of direct mixing of maternal and fetal blood - certain hormones^3,4^, immunoglobulins^5,6^, nutrients^7^, viruses^8^, and toxins^9^ can still pass through this barrier, potentially exposing the fetus to a number of external insults from maternal circulation. Furthermore, those factors in maternal circulation can affect placental function even when they do not cross it^10^, potentially affecting the fetus in spite of the mechanical shield provided by the placenta.

Fetal exposure to factors in maternal circulation provides therefore a plausible avenue for maternal pregnancy exposures to impact the fetus – whereby disruption of maternal homeostasis that arises as a consequence of medical conditions in pregnancy could impact the developmental trajectory of the fetus. Alternatively, genetic variation predisposing women to certain conditions could also confer ASD risk, resulting in familial co-occurrence of ASD and other medical conditions.

To date, some of the maternal diagnoses linked with offspring ASD risk include e.g. depression^11,12^, diabetes^13^, autoimmune diseases^14^, asthma^15^, and recurrent infections^16^. Nevertheless, pregnant women receive many other diagnoses around pregnancy, most of which have not been evaluated for an association with offspring ASD. We have previously shown^17^ that mothers of children with ASD have on average 4 more distinct diagnoses around the pregnancy than mothers of control children - suggesting that the associations between maternal health and ASD may be far more pervasive than just the diagnoses identified to date.

Here we explore the association between offspring ASD and the full spectrum of maternal diagnoses around pregnancy. Using a large, population-based cohort we test the associations between the full range of recorded maternal diagnoses and offspring ASD, applying a series of rigorous screens for potential false positive findings and controlling for the correlation between maternal diagnoses.

## METHODS

### Sample

Our sample is population-based, derived through a case-cohort ascertainment design, as described in detail previously^17,18^. Briefly, the data source was Meuhedet, a large health maintenance organization (HMO) in Israel. Legislation requires all Israeli citizens to purchase medical insurance from a HMO (each offering equivalent medical provision and fees) and prohibits HMOs refusing a citizen membership, thus limiting the risk of ascertainment bias in the sample.

The cohort birth years were defined as January 1^st^ 1997 through December 31^st^ 2008, and children were followed up until January 26^th^ 2015. The study sample comprised of 19.5% of 270,799 children born within the Meuhedet HMO within those years, sampled without stratification, as well as all ASD cases born within those years. Additionally, all siblings of those individuals, born within the cohort birth years, were also included in the sample.

This ascertainment design produced a subcohort including random 19.5% of Meuhedet children born between 1997 and 2008, together will all siblings of ASD cases and controls. Due to random sampling, the subcohort included some ASD cases, and thus overlapped with the individuals selected independently as ASD cases. Family relations (siblings, parents) in the dataset were identified through Meuhedet Family Relations Register.

In order to ensure complete coverage of maternal diagnoses during, and prior to pregnancy (see exposure definition in Maternal diagnosis classification) for all births, we used the subset of children born on, or after January 1^st^ 1999 through December 31^st^ 2008. Due to the low likelihood of reproduction beyond ages 13-55 in females, and below 13 in males, we excluded all children born to mothers younger than 13 or older than 55, or fathers older than 55, as records of those children were likely to include administrative errors.

This study was approved by the institutional review board of the University of Haifa and the Helsinki Ethics Committee of the Meuhedet. Those bodies waived the need for informed consent because the study data were fully de-identified.

### Exposure

The exposures were maternal diagnoses classified according to International Classification of Diseases, Ninth Revision (ICD-9). All diagnostic codes were identified through the Meuhedet Diagnostic Classification Register. The ICD-9 diagnoses are organized in a hierarchical way, with 4 levels of information going from least to most specific to the given diagnosis (see **Fig. 1** for an example). For the current analyses, we used the information at level 3 as it provides both a useful summary, and sufficient detail regarding the underlying health condition. In the current analyses, we included all recorded maternal diagnoses, irrespective of the internal system they affect, their chronicity or severity. The exposure period was defined as the pregnancy period (here defined as 271 days prior to child’s date of birth) and the preceding year (i.e. a total of 636 days prior to child’s date of birth). Widening of the exposure window beyond pregnancy only allowed us to ascertain temporally proximal diagnoses which could still exert effects on the fetus.

**Figure 1.**
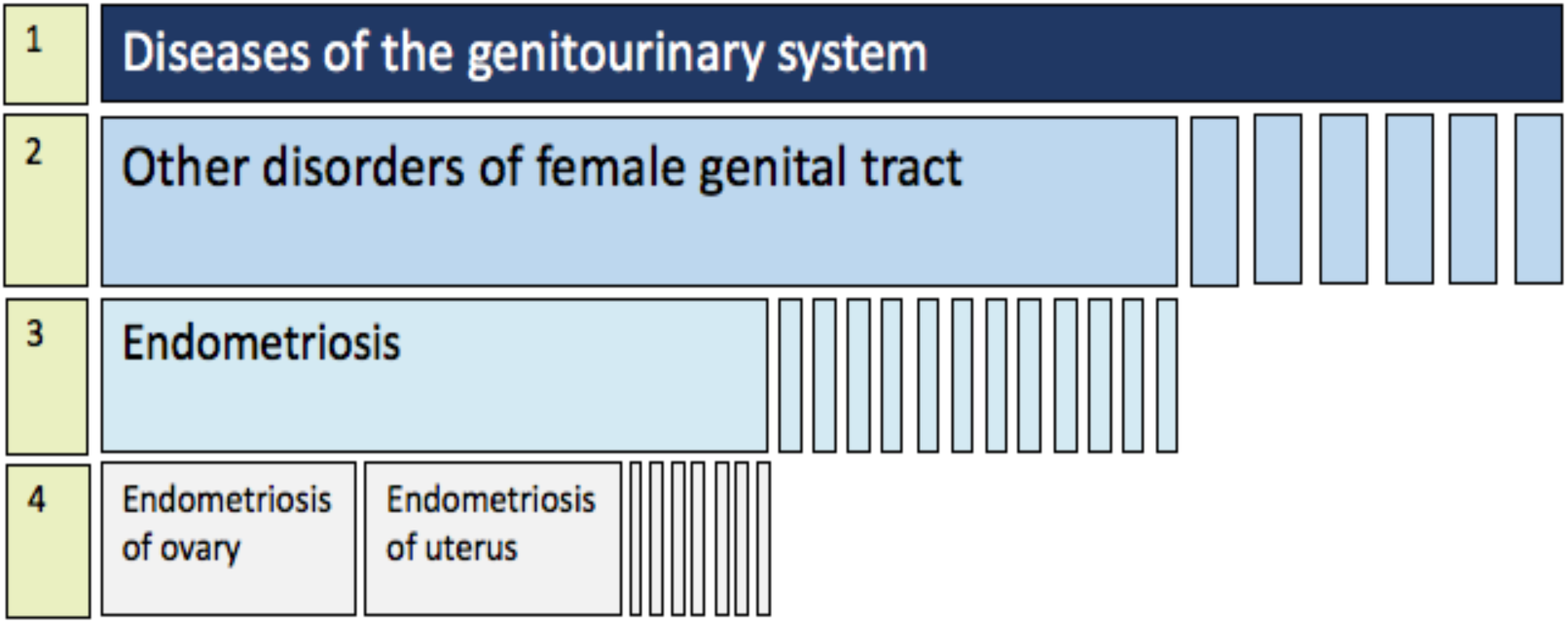
Example of the hierarchical organization of the ICD-9 taxonomy. ICD-9 categories are organized from the most general (level 1, top row), through most specific diagnostic codes (level 4, bottom row). Level 3 diagnoses were used in the current study.

Prior to the analyses, we filtered out the diagnoses not recorded in any women during the exposure period, and those where the available information did not allow assignment to any ICD-9 code. The exposures were coded as binary variables (yes/no) indicating presence/absence of any given level 3, ICD-9 diagnosis within the exposure period. Earlier analyses have demonstrated that the diagnostic rates of all interrogated conditions approximate those recorded in Sweden^17^.

### Outcome

ASD cases were ascertained using the ICD-9 and ICD-10 criteria for ASD from the International Classification of Diseases (ICD-9 codes 299, and ICD-10 codes F84, including all subcodes within those categories). In the Meuhedet system, all children with suspected ASD underwent evaluation by a panel of social workers, a psychologist, and either a trained psychiatrist, a developmental behavioral pediatrician, or a child neurologist. The final diagnosis was made by a board-certified developmental behavioral pediatrician. All children were followed up until January 26, 2015, first ASD diagnosis, or death, whichever occurred first. Earlier analyses have demonstrated that age-specific ASD prevalence in this sample matches that reported previously in Israel^17^.

### Covariates

In all adjusted models the covariates included child’s year of birth, maternal age at child’s birth, number of all maternal diagnoses during the exposure period (636 days prior to child’s date of birth), and residential socioeconomic status (SES). Information about all covariates was obtained from the Meuhedet records except for socioeconomic status (SES), which was obtained from Central Bureau of Statistics Registry^19^. Residential SES was derived from household census data and represented an index of the number of electrical appliances per person and per capita income. In unadjusted models the only covariate was child’s year of birth.

### Statistical analysis

All analyses were done using R software, version 3.6.3^20^. Relative risk of ASD and the associated two-sided 95% confidence intervals were estimated by the hazard ratios (HRs) from Cox proportional hazard regression models using *survival* package^21^. Proportional hazard assumption was examined using standardized Schoenefeld residuals^22^. In all models, we adjusted for correlation between the data, possible due to presence of siblings in the dataset, by calculating robust standard error estimates^23^.

The covariates included: (i) child’s year of birth – to account for the temporal changes in ASD diagnosis ^24^, as well as other possible temporal effects influencing the patterns of maternal diagnoses; (ii) maternal age at child’s birth – to account for differential ASD risk observed across different maternal age categories^25,26^, and the age-dependent nature of many maternal medical diagnoses; (iii) SES – to account for factors that could contribute to differential healthcare utilization, impacting the probabilities of both any maternal diagnosis and ASD^24^; (iv) number of maternal diagnoses in the exposure period – to account for differential utilization of healthcare around pregnancy, which influences the likelihood of maternal receipt of any diagnosis, and is also associated with ASD risk^17,27^.

We applied inverse probability weighting to account for the sample ascertainment procedures, based on both ASD status, and family size (i.e. due to inclusion of siblings of the randomly selected 19.5% individuals, larger families were more likely to be ascertained). Each individual was assigned a weight. All ASD cases and their siblings received a weight of 1. For other control individuals in the subcohort that weight was always higher, reflecting lower probability of being included in the sample for those individuals; the exact weight given to those individuals was dependent on the family size, with individuals from smaller families receiving higher regression weights (see **Table S1**).

#### Filtering of potential false positive findings

Statistical analyses of the associations between maternal diagnoses and ASD risk in offspring was conducted in several steps in order to filter out the statistically significant associations with high likelihood of representing Type I errors.

First, we filtered out maternal diagnoses recorded in fewer than 10 cases and 10 controls. This is due to instability of the maximum likelihood regression coefficients when the number of events per variable is small, a problem known as sparse data bias^28^. Imposing the cut-off of 10 events per variable has been shown as suitable to eliminate such bias^29^.

Threshold for nominal statistical significance for analytical p-values was set at 0.05. For each of the nominally significant diagnoses in Cox proportional hazards model, we performed permutation testing. Such tests allow the estimation of the probability of observing any given association under the null, i.e. in the absence of a real association between exposure and outcome^30^. The rate of ASD, relationship between the follow-up time and ASD status, and correlations between the exposure and covariates, remained the same in the original dataset and under each permutation; however, the recorded associations between ASD and maternal exposure were broken in the permutations – allowing estimation of regression coefficients expected under the null. We performed 1,000 permutations of each nominally significant association to determine empirical P-values (defined as the proportion of tests producing a P-value equal to, or smaller than the analytical P-value (*i.e*. in the non-permuted dataset)). To validate the analytical P-value derived from the analyses in the original dataset, we required that at least 95% of the P-values observed after permutations are higher than that analytical P-value, i.e. an empirical P-value <0.05.

Due to the multitude of tests being performed (equal to the number of distinct maternal diagnoses), we then used false discovery rate correction for multiple testing (Benjamini-Hochberg^31^) on empirical p-values. The permissible false discovery rate (Q-value) was set at 0.05.

Finally, all diagnoses that remained significant after the FDR correction were entered into a joint multiple regression model, in order to account for possible correlation between those diagnoses. The model specifications were analogous to those used in initial adjusted univariate regressions, as described above. An outline of the analytical framework is presented in **Figure S1**.

#### Missing Data and Sensitivity Analyses

In order to evaluate the effects of missing covariate data on the results, we verified consistency of the results in the data with and without missing data imputed using multivariate imputation by chained equation implemented in the *mice* package^32^.

Earlier studies have suggested high levels of comorbidity between ASD and other psychiatric and physical conditions^33^. In order to ensure that the associations observed in our study do not reflect comorbidities in mothers with ASD, we re-analysed the data after exclusion of all families where the mother herself had a recorded ASD diagnosis.

## RESULTS

There were 88,713 children in the Meuhedet dataset born 1999-2008. After removing records with missing values on any of the covariates, we retained 80,202 individuals (no individuals were removed due to missing maternal age or year of birth, but 8,511 individuals had missing SES information). Additionally, 17 individuals were removed due to being born to mothers younger than 13 or higher than 55, or fathers older than 55, producing a final analytical sample of 80,187 children (90% of the 1999-2008 birth cohort), born to 30,864 mothers. There were 1,132 children diagnosed with ASD, and 79,055 controls, with the median age at the end of follow-up 11.2 (IQR: 8.8-13.6). We identified 5 mothers of children in the cohort who themselves received an ASD diagnosis during the follow-up period. Sample characteristics are presented in **Table 1** (see **Table S2** for characteristics of the sample before exclusions).

**Table 1.** Analytical sample demographic characteristics (N=80,193).

|  | Controls<br>(N=79,055) | ASD<br>(N=1,132) |
| --- | --- | --- |
| % female | 49.2 | 20.1 |
| Mean SES (SD) | 7.6 (4.3) | 10.3 (4.2) |
| Mean maternal age (SD) | 29.8 (5.4) | 30.0 (5.4) |
| Mean paternal age (SD) | 32.4 (6.1) | 32.9 (6.2) |
| N children in the dataset born<br>each year (% total sample) |  |  |
| 1999 | 7,991 (10.1%) | 88 (7.8%) |
| 2000 | 8,068 (10.2%) | 91 (8.0%) |
| 2001 | 8,161 (10.3%) | 111 (9.8%) |
| 2002 | 8,431 (10.7%) | 114 (10.1%) |
| 2003 | 8,555 (10.8%) | 145 (12.8%) |
| 2004 | 8,451 (10.7%) | 153 (13.5%) |
| 2005 | 8,162 (10.3%) | 135 (11.9%) |
| 2006 | 7,824 (9.9%) | 141 (12.5%) |
| 2007 | 8,057 (10.2%) | 126 (11.1%) |
| 2008 | 5,335 (6.8%) | 28 (2.5%) |
| % mothers with ASD | <0.1 | 0.3 |

### Associations between maternal diagnoses and offspring ASD

There were 822 distinct level 3 ICD-9 diagnoses recorded in women during pregnancy and the preceding year in the Meuhedet records (**Fig. S2**). Of those, 453 maternal diagnoses were nominally statistically significantly associated with offspring ASD risk (**Fig. S3**). Most of those associations were between ASD and diagnoses recorded in mothers of very few controls and none of the cases, consequently producing associations with low hazard ratios (point estimate HR<0.01; **Fig. S4**). Only 52 (11 %) of those 453 diagnoses were associated with an increased risk of ASD (point estimate HR >1; **Fig. S5**)

After filtering out diagnoses recorded in mothers of less than 10 cases or 10 controls, due to unreliability of the regression coefficients for sparse predictors, we retained 148 maternal diagnoses in the dataset, 22 of which were nominally significantly associated with offspring ASD (**Table 2;** see **Table S3** for all results), distributed across a broad range of diagnostic categories (**Fig. 2a**). Fourteen of those diagnoses (64%) were associated with an increased risk of ASD (HR > 1; *cf*. 42% in unadjusted models (**Table S4**)). Frequencies of those diagnoses in cases and controls are presented in **Table S5**.

**Table 2.** Top associations between maternal diagnoses and offspring ASD, ordered by the p-values from adjusted univariate models. Maternal diagnoses significantly associated with ASD in the final multiple regression are highlighted in blue.

| ICD-9 | Diagnosis / Medical code | HR (CI)* | Analytical p-value* | Empirical p-value* | Multiple regression p-value |
| --- | --- | --- | --- | --- | --- |
| V22 | Supervision of normal pregnancy | 0.62 (0.54 - 0.71) | 1.80E<sup>-11</sup> | 0 | 4.71E<sup>-10</sup> |
| 401 | Essential hypertension | 2.74 (1.92 - 3.90) | 2.43E<sup>-08</sup> | 0 | 3.62E<sup>-05</sup> |
| 110 | Dermatophytosis | 0.57 (0.43 - 0.74) | 3.13E<sup>-05</sup> | 0 | 9.35E<sup>-05</sup> |
| 622 | Noninflammatory disorders of cervix | 1.88 (1.38 - 2.57) | 7.06E<sup>-05</sup> | 0 | 1.82E<sup>-04</sup> |
| V71 | Observation and evaluation for suspected conditions not found | 1.53 (1.23 - 1.91) | 1.66E<sup>-04</sup> | 1.00E<sup>-3</sup> | 5.37E<sup>-04</sup> |
| 642 | Hypertension complicating pregnancy childbirth and the puerperium | 1.77 (1.30 - 2.42) | 2.91E<sup>-04</sup> | 0 | 5.00E<sup>-02</sup> |
| 278 | Overweight, obesity and other hyperalimentation | 1.69 (1.26 - 2.27) | 5.49E<sup>-04</sup> | 2.00E<sup>-3</sup> | 2.00E<sup>-02</sup> |
| 463 | Acute tonsillitis | 0.78 (0.67 - 0.91) | 1.58E<sup>-03</sup> | 2.00E<sup>-3</sup> | 3.90E<sup>-03</sup> |
| 311 | Depressive disorder, not elsewhere classified | 2.11 (1.32 - 3.35) | 1.70E<sup>-03</sup> | 6.00E<sup>-3</sup> | 6.92E<sup>-03</sup> |
| 734 | Flat foot | 2.10 (1.26 - 3.52) | 4.53E<sup>-03</sup> | 4.00E<sup>-3</sup> | 3.28E<sup>-03</sup> |
| 788 | Symptoms involving urinary system | 1.40 (1.09 - 1.80) | 9.47E<sup>-03</sup> | 7.00E<sup>-3</sup> | 1.67E<sup>-02</sup> |
| V68 | Encounters for administrative purposes | 0.76 (0.61 - 0.93) | 9.65E<sup>-03</sup> | 6.00E<sup>-3</sup> | 1.63E<sup>-02</sup> |
| 790 | Nonspecific findings on examination of blood | 1.60 (1.12 - 2.29) | 1.04E<sup>-02</sup> | 1.50E<sup>-2</sup> | 3.89E<sup>-02</sup> |
| 671 | Venous complications in pregnancy and the puerperium | 0.56 (0.36 - 0.88) | 1.19E<sup>-02</sup> | 9.00E<sup>-3</sup> | 2.82E<sup>-02</sup> |
| 564 | Functional digestive disorders not elsewhere classified | 1.41 (1.07 - 1.85) | 1.60E<sup>-02</sup> | 1.80E<sup>-2</sup> | 4.91E<sup>-02</sup> |
| 691 | Atopic dermatitis and related conditions | 0.50 (0.28 - 0.88) | 1.62E<sup>-02</sup> | 2.40E<sup>-2</sup> | 3.54E<sup>-02</sup> |
| 783 | Symptoms concerning nutrition metabolism and development | 1.60 (1.06 - 2.41) | 2.46E<sup>-02</sup> | 3.30E<sup>-2</sup> | 6.71E<sup>-02</sup> |
| 787 | Symptoms involving digestive system | 1.27 (1.03 - 1.56) | 2.58E<sup>-02</sup> | 2.00E<sup>-2</sup> | 7.32E<sup>-02</sup> |
| 628 | Female infertility | 1.22 (1.02 - 1.45) | 3.07E<sup>-02</sup> | 2.90E<sup>-2</sup> | 1.35E<sup>-01</sup> |
| 256 | Ovarian dysfunction | 1.59 (1.03 - 2.44) | 3.58E<sup>-02</sup> | 4.10E<sup>-2</sup> | 2.15E<sup>-01</sup> |
| 692 | Contact dermatitis and other eczema | 0.79 (0.64 - 0.99) | 3.77E<sup>-02</sup> | 3.60E<sup>-2</sup> | 1.46E<sup>-01</sup> |
| 79 | Viral and chlamydial infection in conditions classified elsewhere and of unspecified site | 0.84 (0.70 - 1.00) | 4.62E<sup>-02</sup> | 3.70E<sup>-2</sup> | 6.08E<sup>-02</sup> |

\* univariate models, adjusted for maternal age at child's birth, child's year of birth, SES and total number of diagnoses in pregnancy;

**Figure 2.**
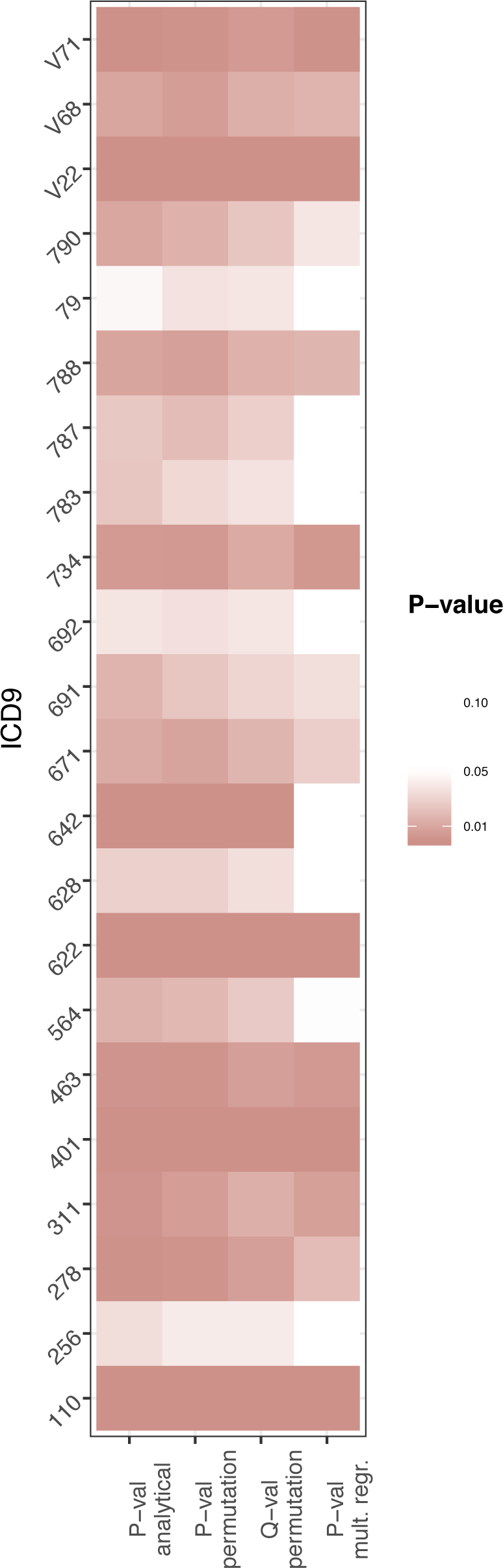
Diagnoses nominally associated with ASD through the consecutive stages of filtering.

**Figure 3.**
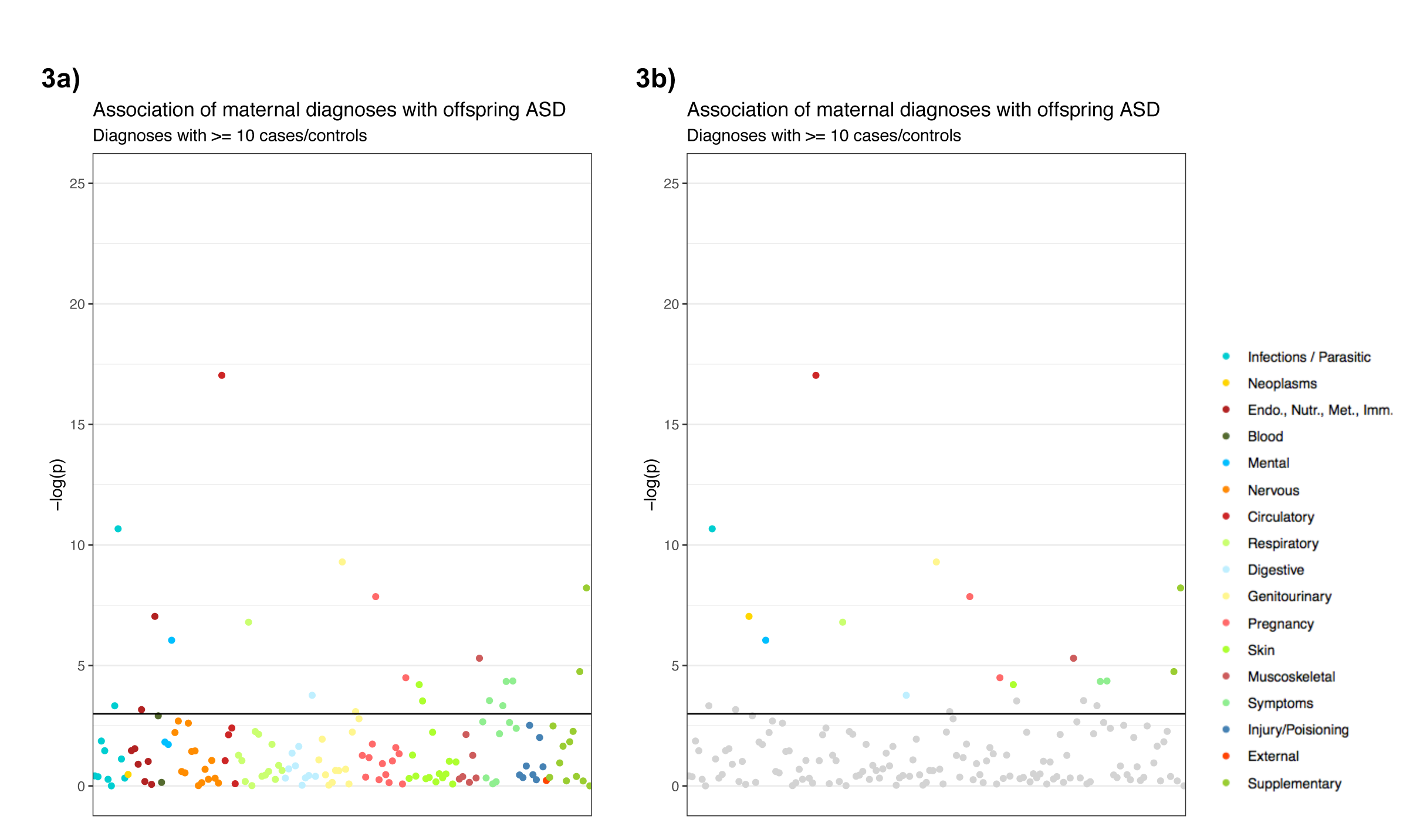
P-values of the associations between maternal diagnoses and ASD, for diagnoses recorded in at least 10 cases and 10 controls. Each dot represents a −log(p-value) of an association between a distinct maternal diagnosis (level 3 of the ICD-9 codes) and offspring ASD. The results are color-coded by broad ICD-9 categories (level 1), as shown in the legend. **Fig. 3A** represents p-values for all diagnoses recorded in at least 10 cases and 10 controls; **Fig 3B** shows the same results, but the diagnoses that were filtered out in permutation testing or subsequent control for multiple testing are shown in grey (N.B. the diagnoses that were non-significant in the multiple regression step are <u>not</u> shown in grey).

Permutation testing revealed that none of those 22 significant associations could arise under the null (empirical p-value < 0.05), and all of them remained significant after controlling for multiple testing. In the final multiple regression step, 16 of those diagnoses remained significantly associated with ASD (**Table 2; Figure 2b**). The Schoenefeld residuals did not indicate violation of the proportional hazards assumption (**Table S6**).

Frequencies of ICD-9 level 3 diagnoses recorded in at least 10 cases and 10 controls categorized by broad (level 1) ICD-9 categories, and count of how many of them remained significant through the consecutive filtering stages is presented in **Table S7**.

### Sensitivity analyses

All nominally statistically significant associations in the full sample remained significant after exclusion of women with recorded ASD diagnosis (**Table S7;** n_mothers_=5 and n_children_=13 were excluded). There was also a close alignment between the effect sizes and levels of significance observed when using models with and without data imputation, suggesting data missingness had no substantial effects on the study results (**Table S8**).

## DISCUSSION

Using our approach, we screened over 820 maternal ICD-9 diagnoses around pregnancy for an association with ASD, and through an iterative process of identifying likely false positive associations, we identified 16 maternal diagnoses with robust evidence for an association with offspring ASD. Those 16 diagnoses that passed through all filtering stages included previously reported ASD risk factors, e.g. depression^11^ and hypertension^34^. In addition, our systematic approach allowed us to identify also novel associations that warrant further attention in ASD research – e.g. noninflammatory disorders of the cervix and urinary symptoms.

The systematic nature of our approach allowed comparisons of risk estimates (HRs) and the strength of the evidence for an association (p-value) across a wide range of diagnoses, using a unified analytical process. The key analytical advantages of such approach are (i) facilitated synthesis of information regarding ASD risk factors pertaining to maternal diagnoses, compared to pooling of many individual studies, each investigating single/few diagnoses, and (ii) limiting false positive findings through explicit acknowledgement of, and addressing the issue of multiple testing.

We emphasize that this rigorous and systematic approach does not strengthen, nor attempts to strengthen, causal inference. While the statistical evidence for the presented associations between maternal diagnoses and offspring ASD is strong, the mechanisms underlying those associations remain to be elucidated. Advancing our knowledge about the etiology of ASD will require careful consideration of potential causal factors underlying those associations, including *e.g*. shared genetic effects (whereby maternal genetic variation transmitted to offspring contributes to the risk of both maternal diagnosis and offspring ASD), maternal use of medication, or socioeconomic factors associated with receiving certain diagnoses. For example, apparent “protective” effects (HRs < 1) observed in our study likely reflect differential utilization of healthcare – whereby women with more medical supervision during pregnancy are less likely to have a child with ASD, but through their frequent medical contacts, being also more likely to receive relatively minor diagnoses, e.g. acute tonsillitis or venous complications of pregnancy (ICD-9 463, 761). In support of this, (i) we observed that mother of children with ASD in this sample were less likely to attend a prenatal care appointment or see a healthcare professional for administrative purposes (ICD-9 V22, V68), and (ii) adjusting for the total number of diagnoses received during the exposure period as well as SES removed mostly the associations with HRs <1. Similarly, the association between maternal flat foot (ICD-9 734) and offspring ASD diagnosis likely reflects shared genetic effects, with flat feet frequently reported in individuals with ASD themselves^35,36^.

Therefore, interpreting our results – and other findings on this topic that rely on registry data – it is important to bear in mind that they do not necessarily reflect causal relationships. In order to better understand the etiology of ASD, identifying the maternal diagnoses associated with an increased risk of the disorder needs to be followed by careful examination of the causal factors driving the association, taking into account genetic and social determinants of health and disease, and other factors associated with receiving a medical diagnosis (e.g. medication). While our study represents a novel approach for analyzing associations between maternal diagnoses around pregnancy and ASD, its limitations need to be acknowledged. Importantly, the definition of the exposure window in our study (pregnancy and one year preceding it) likely did not allow us to ascertain the associations between ASD and maternal chronic diagnoses, particularly those that are well-managed and do not necessitate frequent contacts with healthcare professionals (e.g. diabetes). Therefore, we do not propose that the list of associations detected by us is exhaustive. Likewise, the results need to be interpreted as specific to the sample at hand, and further validation should be sought in other cohorts. Rates of ASD differ by country^37^ – with prevalence in Israel (0.5%^24^) representing a relatively low estimate compared to the US (1.7%^38^) – as likely do patterns of maternal morbidity in pregnancy. Replication of the observed associations in other cohorts will be thus crucial for determination of the robustness of the associations reported here, an essential step prior to causal analysis. Furthermore, exposure in our study was defined as presence of at least 1 record of a given diagnosis during the exposure period, rather than 2, as reported in some other studies (e.g. ^39^). While this could introduce some fraction of false positives (non-exposed children classified as exposed, *e.g*. due to administrative errors) our study focused on relatively common diagnoses where those would have limited influence; additionally, false negatives (exposed children classified as not exposed) – would be more likely in a study with a relatively narrow exposure period, compromising statistical power. Finally, we did not have access to information allowing to estimate family-level SES, and thus could only control for residential measures of SES. While those two are expected to strongly correlate, this correlation will not be perfect, and thus the SES adjustment in our study should be considered incomplete.

In conclusion, our work applied a novel approach for identifying associations between maternal diagnoses around pregnancy and offspring ASD, consisting of systematic testing of a wide range of maternal diagnoses, followed by a series of validation steps to filter out the likely false positive associations. We showed that such an approach replicates many of the previously reported ASD maternal clinical risk factors, and identifies novel associations, warranting replication and follow-up in future studies designed to elucidate the underlying causal mechanisms.

## Data Availability

Data access rules do not permit public sharing of the data. Interested researchers should discuss access options with Arad Kodesh and Stephen Levine.

## ACKNOWLEDGEMENTS

We would like to acknowledge the generous support of the Seaver Foundation.

## Author Contributions

Dr Janecka had full access to all the data in the study and takes responsibility for the integrity of the data and the accuracy of the data analysis.

*Concept and design:* Janecka.

*Acquisition, analysis, or interpretation of data:* Janecka, Kodesh, Levine, Sandin, Reichenberg.

*Drafting of the manuscript:* Janecka, Sandin, Kodesh, Levine, Reichenberg.

*Critical revision of the manuscript for important intellectual content:* Janecka, Kodesh, Levine, Khachadourian, Sandin, Reichenberg, Croen, Rahman, Schlessinger, O’Reilly, Grove, Schendel, Buxbaum.

*Statistical analysis:* Janecka, Levine, Sandin.

*Obtained funding:* Reichenberg, Buxbaum, Sandin, Kodesh, Levine, Schendel.

*Administrative, technical, or material support:* Kodesh, Levine, Sandin, Reichenberg.

